# Lessons from web scraping coroners’ Prevention of Future Deaths reports

**DOI:** 10.1101/2022.10.19.22281083

**Authors:** Qingyang Zhang, Georgia C. Richards

**Affiliations:** Mansfield College, University of Oxford, Mansfield Road, Oxford OX1 3TF, UK; Centre for Evidence-Based Medicine, Nuffield Department of Primary Care Health Sciences, University of Oxford, Woodstock Road, Oxford, OX2 6GG, UK

**Keywords:** Prevention of Future Deaths Reports, PFDs, Preventable Mortality, Coroners, Inquest, Post-mortem, Web scraper, Technology

## Abstract

In England and Wales, coroners are required to write Prevention of Future Deaths reports (PFDs) when a death is deemed preventable so that action is taken to avert similar deaths. Since July 2013, PFDs have been openly available via the Courts and Tribunals Judiciary website (https://www.judiciary.uk/subject/prevention-of-future-deaths/). However, their presentation has been insufficient to identify trends and learn lessons. We designed a web scraper to create the Preventable Deaths Tracker (https://preventabledeathstracker.net/). On 22 June 2022, 4001 PFDs were scraped, analysed, and compared to the Office of National Statistics (ONS) preventable mortality statistics. This commentary summarises the key findings and offers recommendations to improve the PFD system so lessons can be learnt to avert preventable deaths.

---

Since The Coroners Rule 1984,(1) coroners in England and Wales have been required to report and communicate a death when they believe action is needed to prevent similar deaths. These reports, named Prevention of Future Deaths reportsor PFDs (previously Rule 43) are mandated under Paragraph 7 of Schedule 5 of The Coroners and Justice Act 2009,(2) and Regulations 28 and 29 of The Coroners (Investigations) Regulations 2013.(3) Figure 1 shows where a PFD may be written in the coronial process, highlighted using data from the Chief Coroner’s Annual Report.(4)

**Figure 1:**
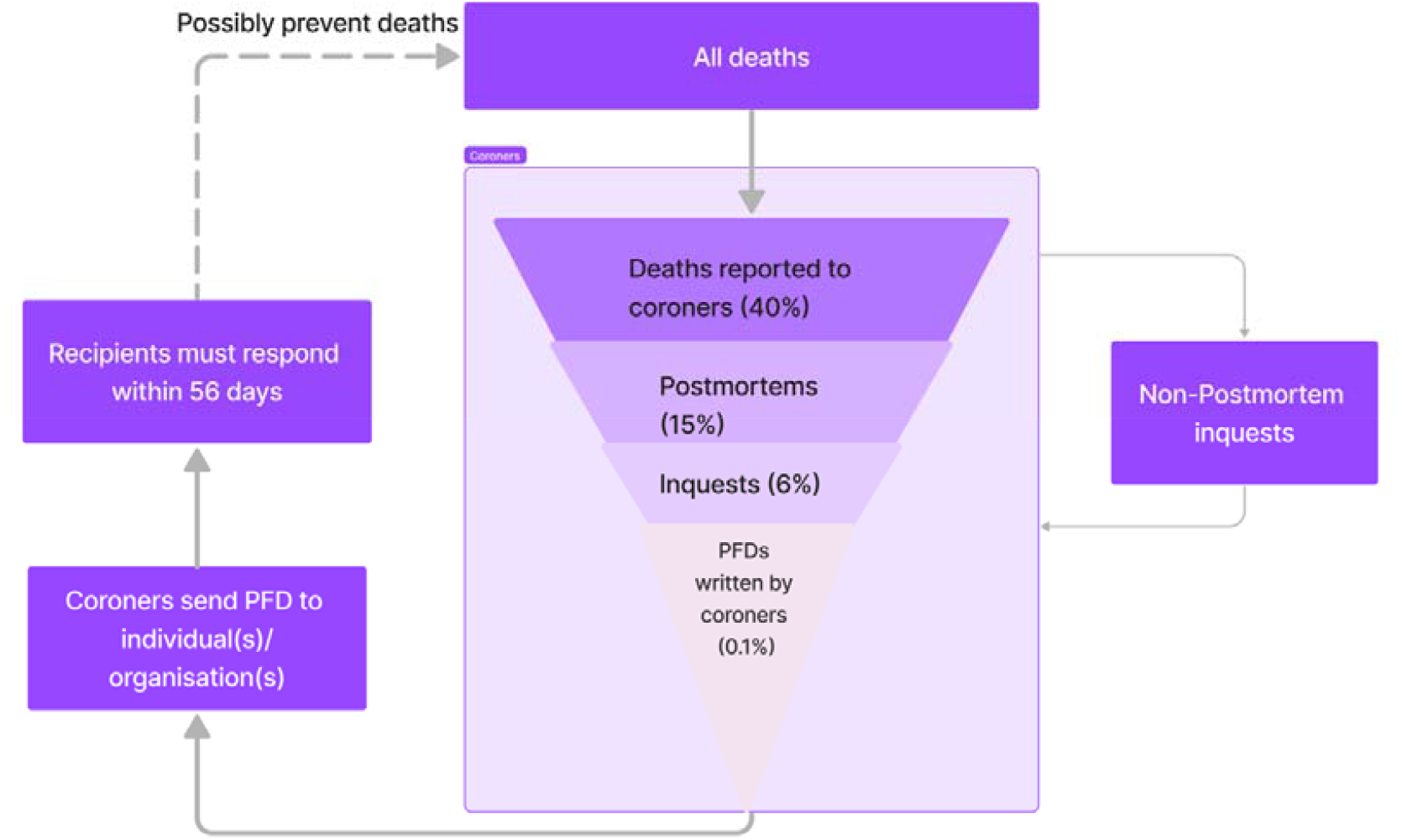
Summary of the Prevention of Future Deaths reports (PFDs)writing process. When a death occurs, it may be reported to a coroner. The coroner may request a post-mortem examination and/or an inquest to investigate the circumstances and causes of death. If the death is deemed preventable, the coroner should write a PFD and send it to an individual(s) or organisation(s) who have the power to act. The recipient must respond to the coroner within 56 days under statue.(3)The PFD and its available response(s)may be published on the Courts and Tribunals Judiciary website, where PFDs may be reviewed to learn lessons to prevent future deaths. However, this process is not audited or monitored to ensure responses are made or that actions are being taken. The percentage of deaths proceeding through each stage was reported in the Chief Coroner’s Annual report for 2019.(4)

PFDs and their responses are openly published on the Courts and Tribunals Judiciary website.(5) We designed a web scraper, which is openly available,(6)(7) to collect PFDs from the Courts and Tribunals Judiciary website and create a searchable database to develop the Preventable Deaths Tracker website.(8) The database is now an open resource that can be used to identify trends and provide important lessons for policy and practice. Previous studies have used the Preventable Deaths Database to examine preventable deaths during the Covid-19 pandemic,(9) cardiovascular disease and anticoagulants,(10) and deaths from medications purchased online.(11)

On 22 June 2022, information from 4001 PFDs were scraped from the Courts and Tribunals Judiciary website and analysed. Here we present insights from this analysis including the types of preventable deaths, geographical variations in the writing and responding to PFDs, and comparisons with the Office of National Statistics (ONS) preventable mortality statistics.(12) All code and data used in this study is openly available on GitHub.(7)

## Types of preventable deaths

When a PFD is published on the Courts and Tribunals Judiciary website, it is assigned to one or more category. As at 22 June 2022, the most common (44%; N=1772) PFD category was “hospital (clinical procedures and medical management)” related deaths (Figure 2). In 2021, there was an increase in police, mental health (including suicide), and product-related deaths(Figure 3). The increase in these categories may be due to the complexities of such deaths reported to coroners, the impact of the Covid-19 pandemic, and improvements in categorising of PFDs. Between 2013 and 2018, the median number of categories assigned to a PFD on the Courts and Tribunals Judiciary website was 0 (IQR: 0-0). In 2021, this statistic improved to a median of 1 (IQR: 0-2). Despite these improvements,73% (N=2939) of deaths are not categorised on the Courts and Tribunals Judiciary website. Most recently in 2021, 33% (n=144/433) of deaths were not categorised.

**Figure 2:**
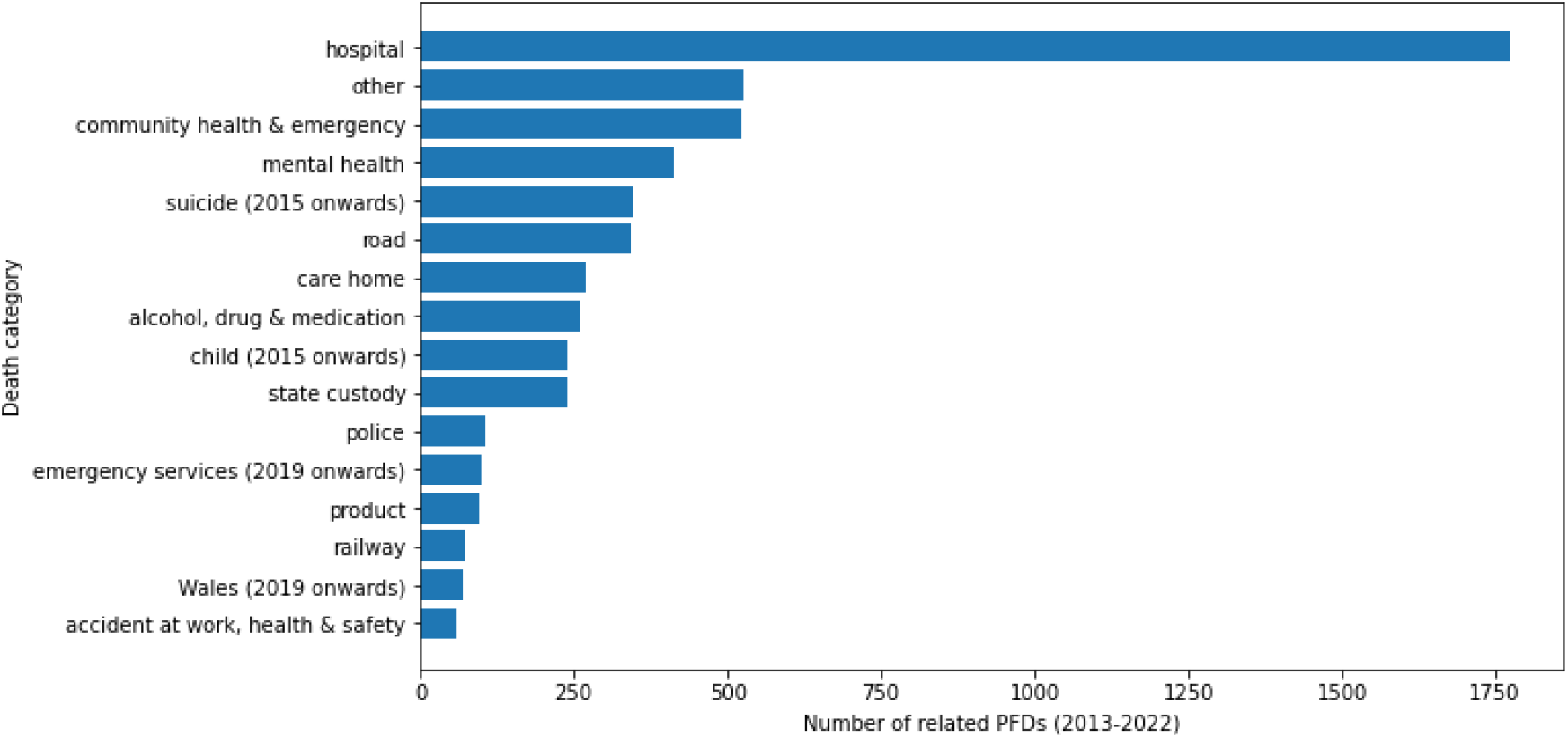
Frequency of Prevention of Future Deaths reports (PFDs)written between July 2013 and 22 June 2022 by coroners in England and Wales, grouped by death categories. Each PFD is assigned to one or more categories on the Courts and Tribunals Judiciary website. As of 22 June 2022, there were 16 categories.

**Figure 3:**
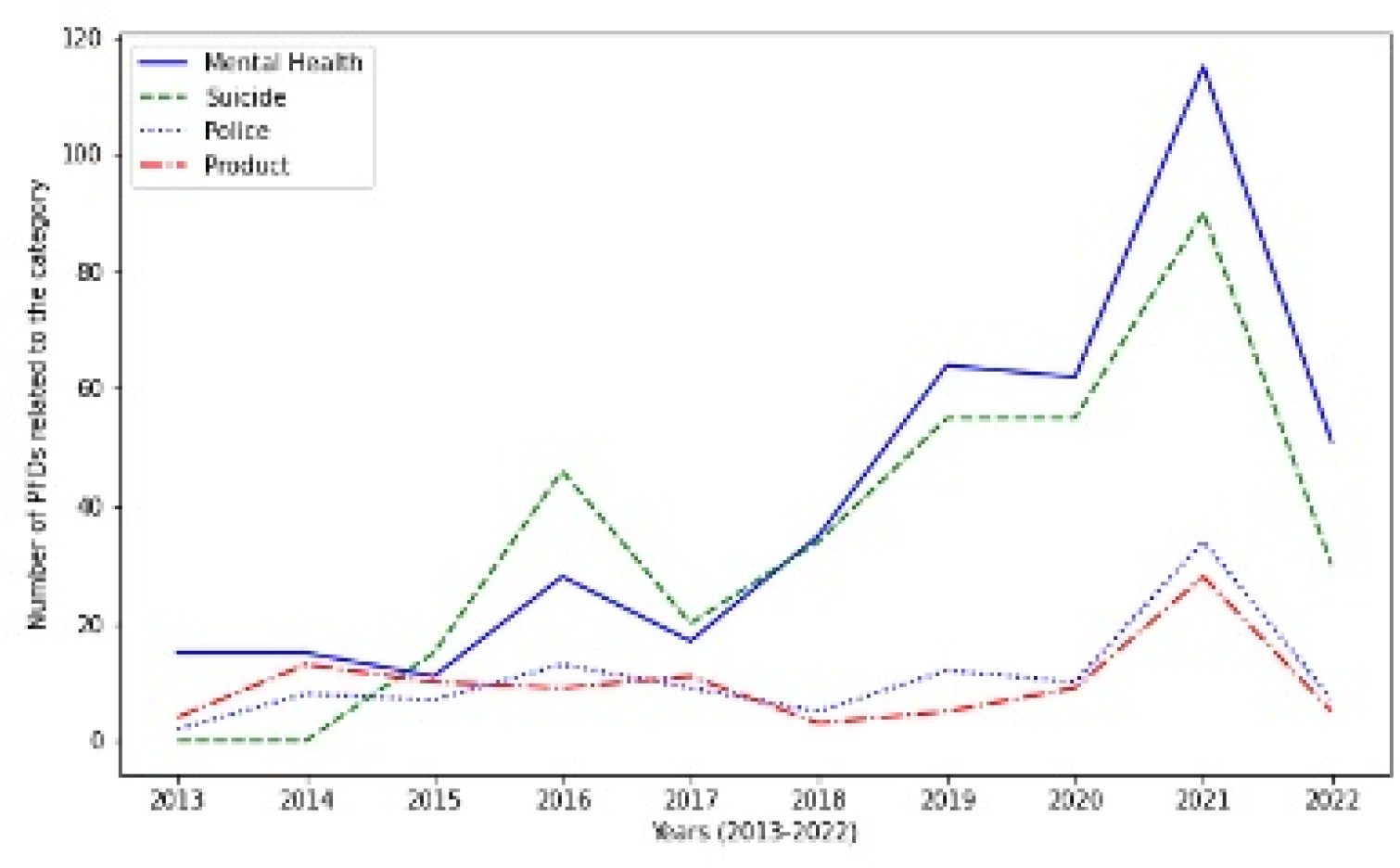
Prevention of Future Deaths reports (PFDs) written by coroners in England and Walesbetween July 2013 and 22 June 2022 that were assigned to categorieson the Courts and Tribunals Judiciary website, which increased in 2021, including mental health related (blue), suicide (orange), police related (green) and product related (red) deaths.

Categories were not consistently defined overtime. The Courts and Tribunals Judiciary website introduced “Child deaths” and “Suicide related deaths” in 2015 and “Emergency services related deaths” and “Wales prevention of future deaths report” in 2019. After the introduction of the “Emergency services related deaths”, the existing category “Community health care and emergency services related deaths” was not modified to exclude emergency services related deaths. This creates ambiguity and uncertainty in the types of preventable deaths. There is also no guidance or criteria that highlights the process used to categorise PFDs. There are internationally recognised classification systems (e.g. the World Health Organisation (WHO) International Classification of Diseases 11^th^ Revision (ICD-11)(13)), which are used by the ONS that could be used by the Courts and Tribunals Judiciary website to improve the quality and ability of PFDs to be analysed and compared to national and international statistics. For further insights on our analyses, visit: https://preventabledeathstracker.net/database/death-categories/.

## Geographical variations in writing PFDs

The median number of PFDs written in each coroner area was 39 (IQR: 17-61 PFDs) and coroners wrote a median of 3 PFDs (IQR: 1-10). Twenty coroners (of 450, top 4.4%) wrote 30% (N=1187) of all PFDs. The most PFDs were written in Manchester South (8%; N=326) and Inner North London (5%; N=208) (Figure 4). However, we could not control for the number of coroners working in each area because of a lack of standardisation and poor data management on the Courts and Tribunals Judiciary website. We identified over 450 coroners that had naming issues, including misspellings, inconsistent use of middle names, varying abbreviations, and variable use of titles and their formatting. Similar patterns were found for date formats (e.g. “November” as “Nevember” (19)) and coroner areas (e.g. “North Yorkshire and York including North Yorkshire Western District”(20)vs. “North Yorkshire (Western)”(21)).

**Figure 4:**
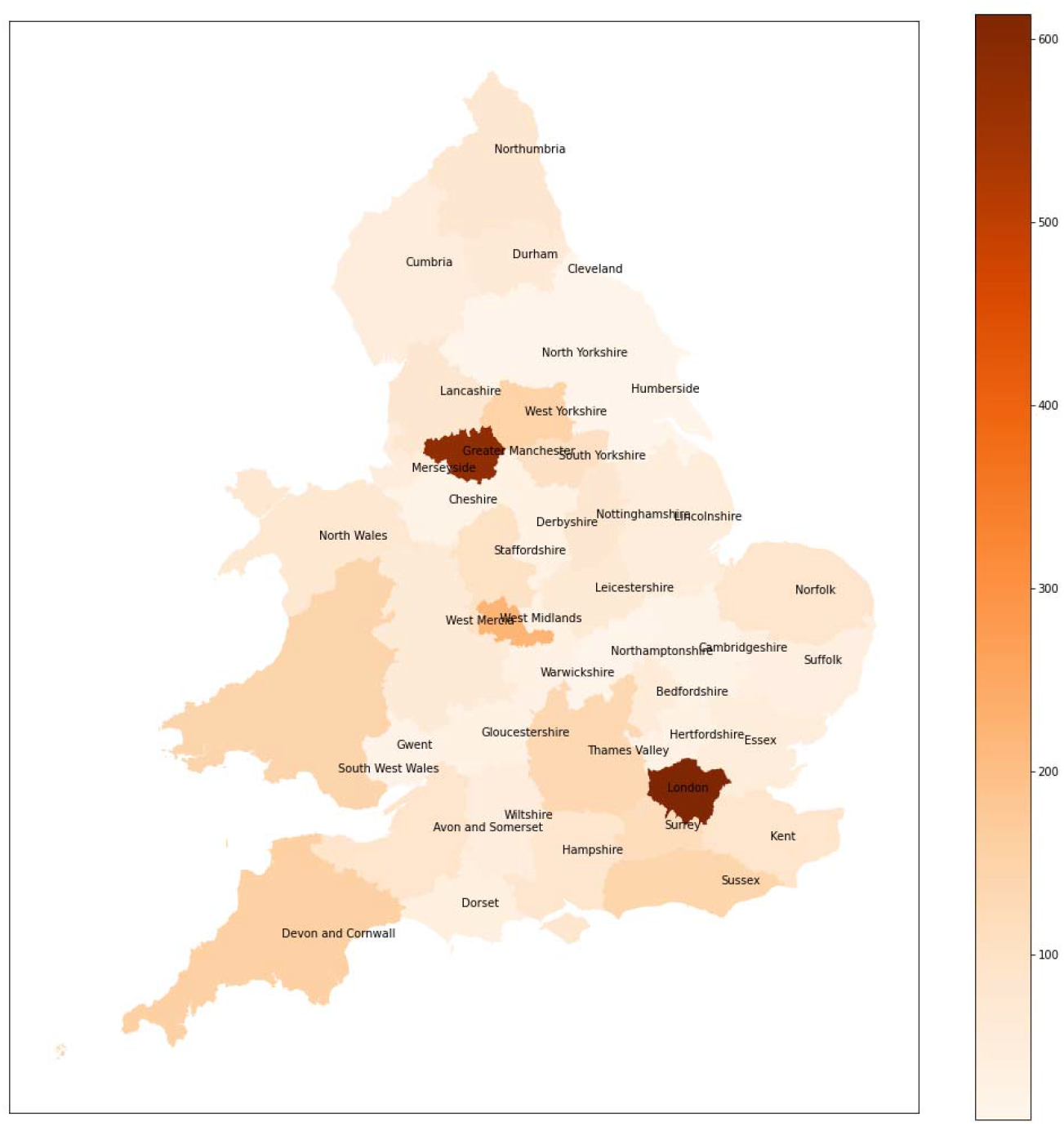
Geographical distribution of Prevention of Future Deaths reports (PFDs) written by coroners in England and Walesbetween July 2013 and 22 June 2022. Due to the lack of coroner area geographical data, the map is plotted using the 42 police areas instead of 85 coroner areas.

Technologies and systems should be implemented to remove such issues, including the use of electronic PFD forms where drop-down boxes provide formatting. Providing the geographical data for coroner areas as a GeoJSON file would also improve data visualisation, as we had to merge coroner areas into the 42 police areas to plot geographical distributions. For further insights on our analyses, visit: https://preventabledeathstracker.net/database/geographical-variation/ and https://preventabledeathstracker.net/database/coroner-names/.

## Response rates to PFDs

PFDs are sent by coroners to individuals or organisations that have the power to act to prevent future deaths. Under law, these individuals or organisations must respond within 56 days.(3) Their responses are also uploaded to the Courts and Tribunals Judiciary website and we used the web scraper to count the number of responses available. As of 22nd June 2022, approximately one in three PFDs (36%; N=1412) did not have an available response and thus whether actions were taken to prevent such deaths is unknown. Sharing such information could help similar organisations or individuals implement changes and learn lessons to reduce preventable deaths.

There is also significant missing information and formatting issues on the Courts and Tribunals Judiciary website that must be considered. More than one quarter (n=1068) of PFDs, mostly between 2013 and 2015, did not list any recipients on the webpage. Six hundred recipients had unstandardised punctuation (e.g. “Kent and Medway NHS Social Care Partnership Trust” and “Kent County Council – Adult Social Care”(14)), which makes it difficult to analyse unless natural language processing is used. For further insights on our analyses, visit: https://preventabledeathstracker.net/database/responses/.

## ONS data vs PFDs

We compared data from PFDs with the Office of National Statistics (ONS) preventable mortality statistics.(12)We found that preventable mortality was higher in Wales than England, despite England having a higher proportion of PFDs written (Table 1).

**Table 1:** Mortality statistics from the Office of National Statistics (ONS) and Prevention of Future Deaths reports (PFDs) grouped by country (England and Wales) between 2014 and 2020.

| **Variable** | **England** | **Wales** |
| --- | --- | --- |
| Total ONS mortality per 100,000 | 908 | 1078 |
| ONS preventable mortality per 100,000 | 149 | 170 |
| PFDs/population per 100,000 | 0.77 | 0.66 |
| Coroners/population per 100,000 | 0.76 | 1.32 |
| PFDs/coroners | 8.71 | 5.91 |

## Conclusions

Our web scraper and the Preventable Deaths Tracker (https://preventabledeathstracker.net/) makes important information accessible for all. Using reproducible data collection methods, we demonstrate that preventable deaths in England and Wales occur mostly in hospitals and that there is wide geographical and coronial variation in the writing and responding to PFDs. However, our analyses alsoreveal areas for improvement, including the need for an internationally recognised system to classify types of deaths and technologies to remove misspelling and formatting inconsistencies, and improve reporting of essential information such as gender and age at death. One third of PFDs were without responses, which illustrates poor compliance with Regulations 29 of The Coroners (Investigations) Regulations 2013. Technologies could also be used to audit compliance with such regulations to ensure that actions are being taken to prevent future deaths. Overall, PFDs hold a rich source of information that should be used by decision-makers to improve public and patient safety.

## Data Availability

All data and code used in this study are openly available on GitHub

https://github.com/georgiarichards/georgiarichards.github.io

